# Non-invasive fasciculation assessment of gastrocnemius failed to show diagnostic or prognostic utility in ALS

**DOI:** 10.1101/2024.04.29.24306549

**Authors:** Robbie Muir, Brendan O’Shea, Raquel Iniesta, Urooba Masood, Cristina Cabassi, Domen Planinc, Emma Hodson-Tole, Emmanuel Drakakis, Martyn Boutelle, Christopher E Shaw, James Bashford

## Abstract

**Objectives:** Amyotrophic lateral sclerosis (ALS) is a neurodegenerative disorder, affecting both upper and lower motor neurons. High-density surface electromyography is a non-invasive measure of muscle fasciculations, a phenomenon prevalent early in the disease. Previous studies highlighted the clinical importance of these measures from the biceps brachii muscle. Our study aimed to investigate the diagnostic and prognostic value of the medial gastrocnemius muscle.

**Methods:** We performed a retrospective study of 49 ALS and 25 control participants. Means comparisons, logistic regression, receiver operating characteristic analysis, Kaplan-Meier analysis, and a multilevel Cox model were used to determine the predictive utility of fasciculation potential (FP) parameters including FP frequency, median amplitude, and amplitude dispersion.

**Results:** FP parameters demonstrated a poor ability to differentiate between ALS and controls, with ROC analysis producing areas under the curve between 0.58 and 0.63. Furthermore, there was no association between FP parameters and ALS prognosis in the Cox model.

**Conclusions:** FP parameters from the medial gastrocnemius muscle are not useful for the diagnosis or prognosis of ALS patients.

**Significance:** Our results highlight the poor clinical utility of fasciculation quantification in the medial gastrocnemius muscle. Future studies should focus on recordings from more clinically relevant muscles earlier in the disease.

**Highlights:**

- Fasciculation parameters from the medial gastrocnemius muscle have poor ability to differentiate ALS patients from controls.
- Fasciculation parameters from the medial gastrocnemius muscle cannot reliably predict the prognosis of ALS patients.
- Future studies should measure more clinically relevant muscles at an earlier stage in the disease course over a protracted period.

## 1. Introduction

Amyotrophic lateral sclerosis (ALS) is a chronic progressive paralytic disorder, characterised by the degeneration of both upper motor neurons (UMNs) and lower motor neurons (LMNs) (Brown and Al-Chalabi, 2017). The ultimate consequence of the condition is progressive muscle weakness leading to death, usually from respiratory failure, with a median survival time of approximately 19 months from diagnosis and 30 months from symptom onset (Gordon, 2013). However, due to disease heterogeneity and several clinical subtypes, it is difficult to predict individual patient survival early in the disease (Van Es et al., 2017).

It is estimated that there are 5000 people living with ALS in the UK at any one time (Opie-Martin et al., 2021), its prevalence higher in men than in women and its incidence increasing with age (Gordon, 2013). However, the onset of ALS is often non-specific and mimics several more common, benign conditions (Van Es et al., 2017). To make a diagnosis, clinicians rely on guidelines such as the Gold Coast criteria (Hannaford et al., 2021), taking a median of 11.5 months after symptom onset (Paganoni et al., 2014) to reach a diagnosis, a third of patients’ life expectancy (Van Es et al., 2017). As a result, in recent years there has been much interest garnered towards identifying the pre-symptomatic stage of the disease and stratifying patients with similar life expectancies. The benefit of such developments would be two-fold. Firstly, by grouping patients into similar clinical subtypes, it could allow for the earlier identification of ALS patients. In turn, this would enhance their recruitment into clinical trials when more motor neurons remain viable (De Carvalho et al., 2008), and therapeutic intervention may be more likely to prevent or delay disease progression (Benatar et al., 2019). Secondly, the prognosis of ALS varies greatly and through the development of a reliable prognostic model, patients could take greater control of their lives and make better plans for their future.

There are several avenues for research, including genetic markers, fluid biomarkers, and neurophysiological techniques (Van Es et al., 2017). In particular, high-density surface electromyography (HDSEMG) provides a promising option as a non-invasive method of recording fasciculations, capable of revealing LMN involvement in body regions otherwise regarded as unaffected (De Carvalho et al., 2008). Fasciculations are spontaneous, involuntary muscle twitches produced by active motor units (MUs), detected as fasciculation potentials (FPs) on electromyography (EMG) (Drost et al., 2007). They are widespread in ALS and can occur in multiple different muscles (Liu et al., 2021) but can also occur in ALS-mimicking disorders and healthy individuals, making their diagnostic utility uncertain (Noto et al., 2018). FPs are frequently the first abnormality recorded by EMG during neurogenic change in ALS (de Carvalho and Swash, 2013), and their importance in its diagnosis has been increasingly recognised, forming a key component of the Gold Coast criteria (Hannaford et al., 2021). While present in healthy individuals and ALS-mimicking disorders, they are shown to have an increased prevalence and specific morphology in ALS (Bashford et al., 2020c). This may be explained by the underlying pathophysiology of the disease, where denervated muscle fibres are reinnervated by surrounding, healthy MUs through the process of chronic partial reinnervation, allowing strength to be maintained early in the disease course (Bashford et al., 2020c).

A previous HDSEMG study by our group showed that FP frequency from biceps brachii (BB) is a valuable predictive marker of ALS, demonstrating an area under a receiver operating characteristic (ROC) analysis curve of 0.89 (95% confidence interval (CI) 0.81-0.98). Chi squared automatic interaction detection (CHAID) decision tree analysis also confirmed FP frequency to be the single best diagnostic measure, showing an 85.9% predictive accuracy (95% CI 75-93.4%) (Tamborska et al., 2020). Prognostically, previous research has shown that FP frequency increases predominately in the early stage of the disease course (Bashford et al., 2020c, Tsugawa et al., 2018), and the rate of change of this process is associated with a higher disease burden (Wannop et al., 2021, Avidan et al., 2021). However, studies have shown that the frequency and amplitude of FPs may fluctuate as the disease progresses (Bashford et al., 2020c), raising the question to its utility as a diagnostic and prognostic tool.

Given the evidence provided by our group for the diagnostic and prognostic utility of FP parameters recorded by HDSEMG in the BB muscle (Tamborska et al., 2020, Wannop et al., 2021), the aim of this study was to explore whether these same parameters were equally insightful in the medial gastrocnemius (MG) muscle among 49 ALS patients and 25 control participants.

## 2. Methods

### 2.1 Study design

This study is a retrospective data analysis, using data from three observational studies undertaken at King’s College Hospital, London, UK. Using the clinical parameters of the patients with ALS, a prospective dataset containing predicted survival length was obtained using the European Network to Cure ALS (ENCALS) Survival Model (Westeneng et al., 2018).

### 2.2 Ethical approval

Ethical approval was obtained from Research Ethics Services in the North of Scotland (ref: 15/NS/0103), East Midlands (ref: 17/EM/0221), and Yorkshire and the Humber (ref: 19/YH/0164).

### 2.3 Participants

Patients attending the motor nerve clinic at KCH between January 2016 and January 2022 were invited to take part in the studies. These included males and females aged 18-80 who were ambulatory and able to give informed consent. Patients were recruited into two groups: ALS (n=49), and neurological controls (n=8) (Table 1). Those recruited into the ALS group were deemed eligible if they were diagnosed with possible, probable, or definite ALS according to the El Escorial criteria within 24 months of symptom onset. Criteria for exclusion included those with atypical or regionally restricted ALS and those with advanced ALS (wheelchair-bound or dependent on non-invasive ventilation). Patients with bulbar onset ALS were required to demonstrate objective involvement in at least one limb.

**Table 1:**
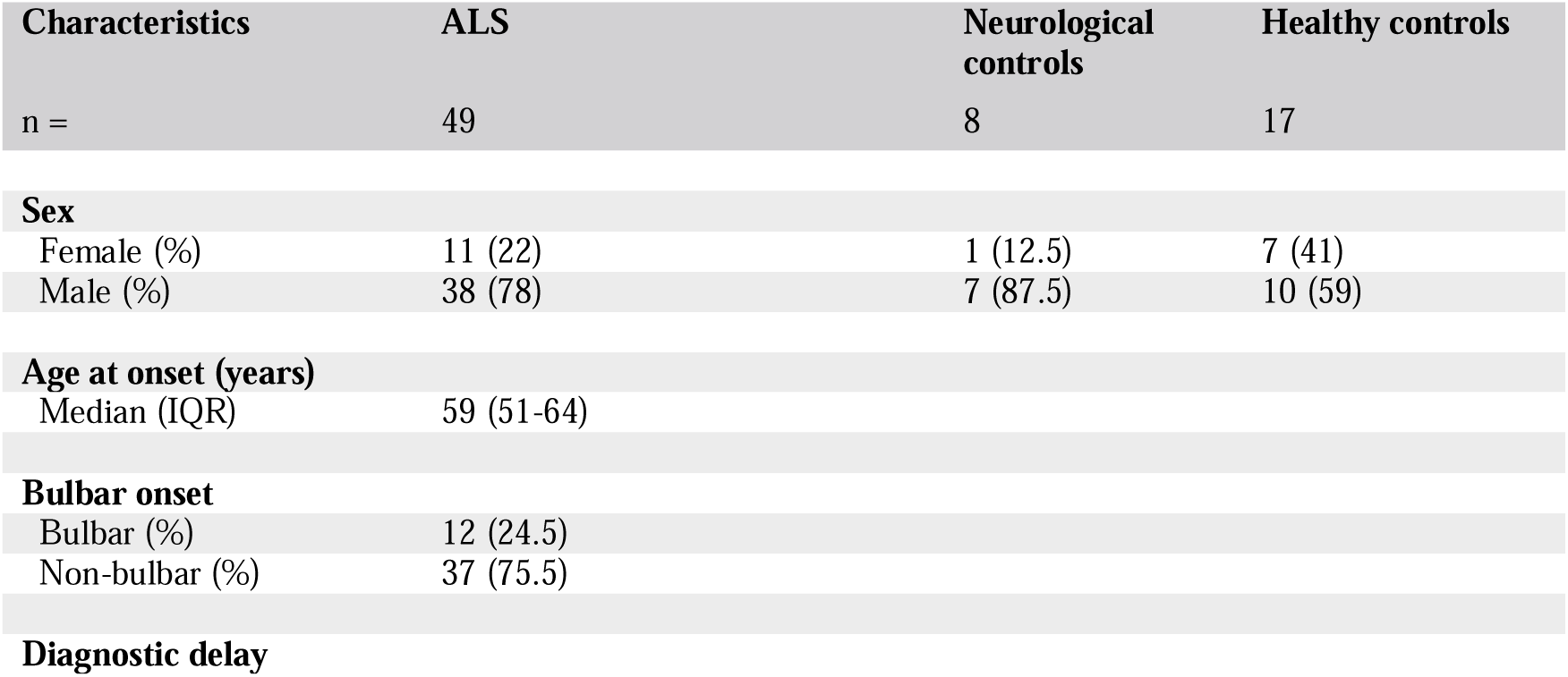
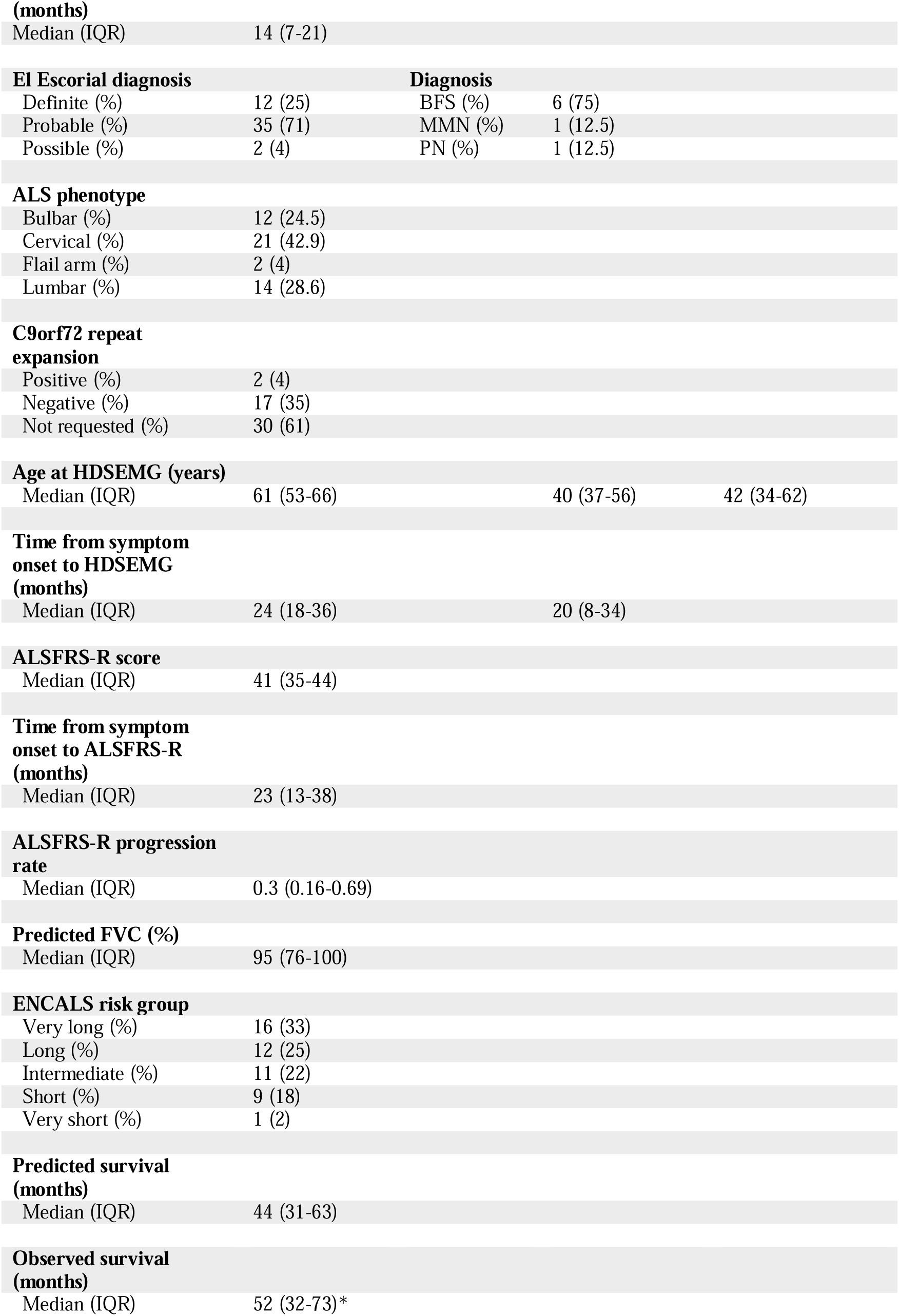
Participant characteristics. Values rounded to the nearest integer where appropriate. *Observed survival data was recorded for 21 patients, with 28 surviving patients censored from the analysis. Abbreviations: ALS, amyotrophic lateral sclerosis; IQR, interquartile range; BFS, benign fasciculation syndrome; MMN, multifocal motor neuropathy; PN, peripheral neuropathy; HDSEMG, high-density surface electromyography; ALSFRS-R, amyotrophic lateral sclerosis functional rating scale revised; FVC, forced vital capacity; ENCALS, The European Network to Cure ALS.

Those recruited into the neurological control group were deemed eligible if they were diagnosed with an ALS-mimicking disorder by a clinician with an expertise in motor neuron disease. The main criterion for exclusion was diagnostic uncertainty. This group consisted of patients with benign fasciculation syndrome (BFS), peripheral neuropathy (PN), and multifocal motor neuropathy (MMN). Electronic health records of these individuals were inspected at a minimum of two years from recruitment, and none had subsequently developed ALS.

The healthy control group (n=17) was recruited from Manchester Metropolitan University between May and September 2019 *via* poster adverts. This group consisted of males and females aged 18-80 who were deemed eligible if they were ambulatory, able to give informed consent, and had no self-reported history of any neurological condition.

### 2.4 Clinical assessment

All patients’ demographic information, including site and date of first symptoms, were recorded where possible. All participants within the ALS and neurological control groups underwent baseline neurological examination, and ALS patients were subsequently classified into the following phenotypes: i) bulbar-onset, ii) cervical-onset, iii) flail arm, iv) lumbar-onset. The disease severity of all participants was determined using the revised amyotrophic lateral sclerosis functional rating scale (ALSFRS-R) at the time of assessment.

### 2.5 HDSEMG recording

Participants underwent either 15- or 30-minute HDSEMG recordings of the BB and MG muscles unilaterally or bilaterally, depending on the original study protocol. The adhesive HDSEMG sensor (comprised of 64 electrodes, arranged in an 8 x 8 grid, electrode diameter 4.5mm, inter-electrode distance 8.5mm; TMS International BV, The Netherlands) was applied centrally over the muscle while the patient was asked to relax in a semi-recumbent position. A reference electrode (3 x 5 cm) was located over the olecranon (when recording BB) and over the dorsum of the foot (when recording MG). Signal amplification was carried out using the Refa-64 Recording System (TMS International, BV, Netherlands), and the raw HDSEMG data were stored as a Polybench file at 2048Hz/channel and processed using the novel surface potential quantification engine (SPiQE) tool (Bashford et al., 2019).

### 2.6 Fasciculation potential parameters

Of all the summary parameters derived from the HDSEMG data by SPiQE, we focused on the following three for the purpose of this study: FP frequency, FP median amplitude, and FP amplitude dispersion. FP frequency is defined as the number of FPs divided by minutes of analysed recording (FP/min). FP median amplitude is defined as the median peak-peak voltage of all FPs detected in each recording (µV). Finally, FP amplitude dispersion is defined as the interquartile range (IQR) of all FP amplitudes detected in each recording (µV).

### 2.7 Statistical analysis

Participant characteristics were summarised using descriptive statistics for all groups. Where bilateral recordings were taken, the values from the side on which the maximal values were recorded were included in the analysis.

#### 2.7.1 Diagnostic

All values were transformed to log(10) to satisfy normality for the statistical tests and the normal distribution, and homogeneity of variance of the transformed values were investigated using Shapiro-Wilk normality tests, inspection of Q-Q plots, and Levene’s tests. Mean comparisons of the FP parameters between groups was carried out using Welch’s one-way analysis of variance (ANOVA) with the Games-Howell *post hoc* test. To assess the association of FP parameters with the diagnosis of ALS, we dichotomised the outcome by combining the neurological and healthy control groups (1= ALS, 0 = not ALS), and performed univariate binary logistic regression, followed by ROC analysis for the assessment of diagnostic utility. To further explore the diagnostic utility, we carried out a CHAID decision tree classification model with 3-fold internal cross-validation, using each of the FP parameters as independent variables. Unless stated otherwise, values were rounded to the nearest third decimal for p-values and the nearest second decimal for remaining values. The threshold for statistical significance was set to p < 0.05 and all statistical analyses were carried out using IBM SPSS Statistics software (version 28, IBM Corp, Armonk, NY, USA).

#### 2.7.2 Prognostic

Prognostic analysis was performed using R version 4.1.1 (R Foundation for Statistical Computing, Vienna, Austria). There were two datasets used in our analysis.

Our first analysis used a prospective survival prediction from a cohort of patients using the ENCALS Survival Model. This prediction was based on each patient’s disease characteristics at their first assessment. Using the same patient cohort in our second analysis, we followed all patients over time and in those patients who died, we recorded the observed survival time for each patient. Those who were still alive at the recording cut-off point, 22^nd^ February 2022, were included as censored in our survival analysis.

A Kaplan-Meier analysis was used to identify the most statistically significant parameters, which were then included in a Cox proportional hazards model. A multilevel Cox model was chosen to evaluate the relationship between muscle FP parameters and disease prognosis, while accounting for the presence of bilateral muscle recordings. This was performed for both predicted and observed survival times. Both Cox models were then evaluated using the cox.zph() function, which showed no violations of the proportional hazards assumption.

## 3. Results

### 3.1 Participant characteristics

### 3.2 Diagnostic

#### 3.2.1 Comparison of fasciculation potential parameters from medial gastrocnemius between groups

Welch’s ANOVA with Games-Howell *post-hoc* test revealed no statistically significant differences between the groups for each of the FP parameters. The results of these analyses are summarised in *Fig. 1* and *Table 2*.

**Fig. 1:**
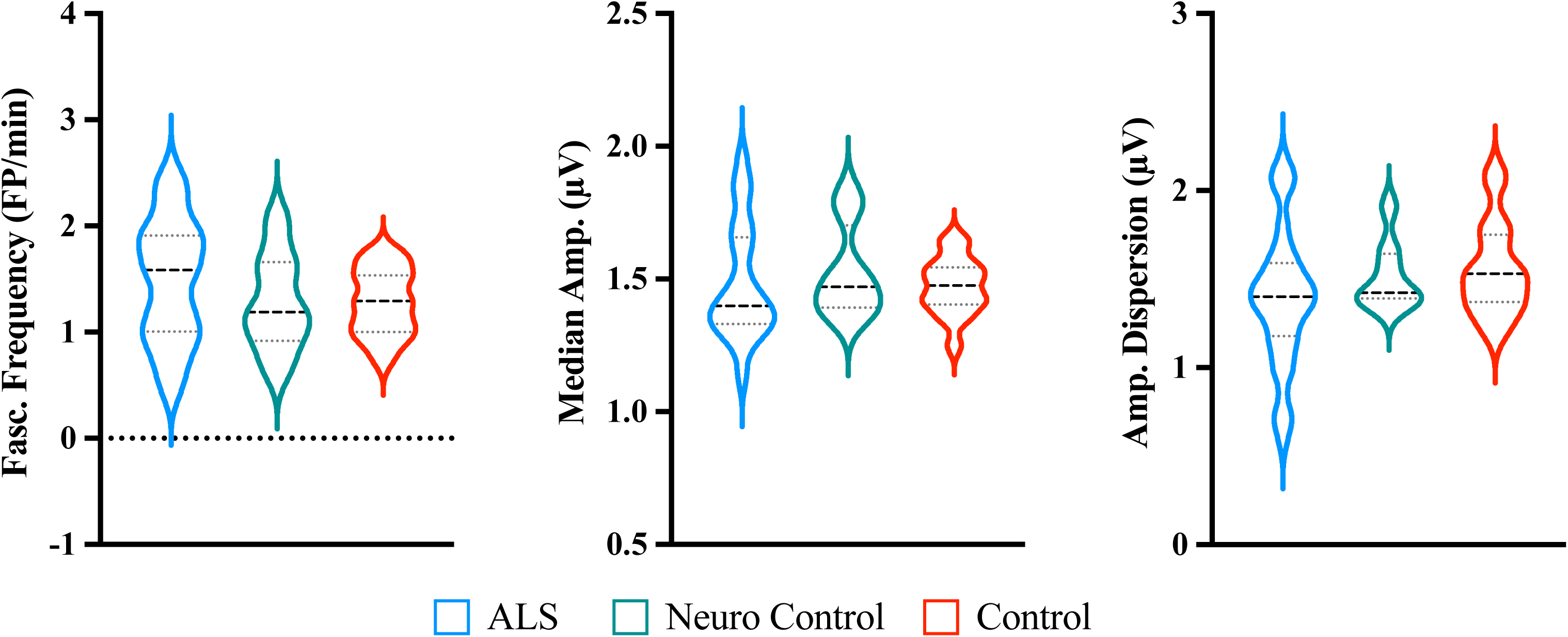
Comparison of fasciculation potential parameters from medial gastrocnemius between the groups. All values were transformed to log(10). Welch’s ANOVA with Games-Howell post-hoc test revealed no statistically significant difference between the means of the groups for each of the fasciculation potential parameters. Abbreviations: FP, fasciculation potential; ALS, amyotrophic lateral sclerosis.

**Table 2:** Comparison of fasciculation parameters from medial gastrocnemius between groups. *FP median amplitude data missing from 1 participant (n=48). ^†^FP amplitude dispersion data missing from 1 participant (n=48). Abbreviations: ALS, amyotrophic lateral sclerosis; FP, fasciculation potential; min, minute; IQR, interquartile range; ANOVA, analysis of variance.

| Fasciculation parameter<br>n = | ALS<br>49 | Neurological controls<br>8 | Healthy controls<br>17 | Statistical significance |
| --- | --- | --- | --- | --- |
| **FP frequency (FP/min)**<br>Median (IQR) | 31 (8.25-92.35) | 13.08 (6.54-35.41) | 14.65 (8.55-33.8) | Welch's ANOVA<br>$W_{2, 19.21} = 1.71$<br>$p = 0.207$ |
| **FP median amplitude ($\mu V$)**<br>Median (IQR) | 23.6 (20.2-41.85)* | 26.6 (21-39.83) | 28.8 (24.95-34.95) | Welch's ANOVA<br>$W_{2, 19.08} = 0.28$<br>$p = 0.757$ |
| **FP amplitude dispersion ($\mu V$)**<br>Median (IQR) | 21 (11.08-31.43)<sup>†</sup> | 24.95 (20.5-31.3) | 33.5 (22.3-56.5) | Welch's ANOVA<br>$W_{2, 23.82} = 2.00$<br>$p = 0.157$ |

#### 3.2.2 Association of fasciculation potential parameters with the diagnosis of ALS

In the univariate binary logistic regression, none of the FP parameters were significantly associated with the diagnosis of ALS (FP frequency, p = 0.117; FP median amplitude, p = 0.910; FP amplitude dispersion, p = 0.083).

#### 3.2.3 Assessment of diagnostic utility

We further explored the diagnostic utility of these FP parameters by carrying out a ROC analysis (*Fig. 2*). FP frequency, FP median amplitude and FP amplitude dispersion displayed areas under the curve (AUCs) of 0.61 (95% CI 0.48-0.74, p = 0.134), 0.58 (95% CI 0.44-0.71, p = 0.299), and 0.63 (95% CI 0.50-0.76, p = 0.073) respectively, each indicative of poor diagnostic instruments. Lastly, we carried out a CHAID decision tree classification model of ALS, using the FP parameters from the MG alone as independent variables. Due to their redundancy, none of the MG FP parameters were included as independent variables in the model.

**Fig. 2:**
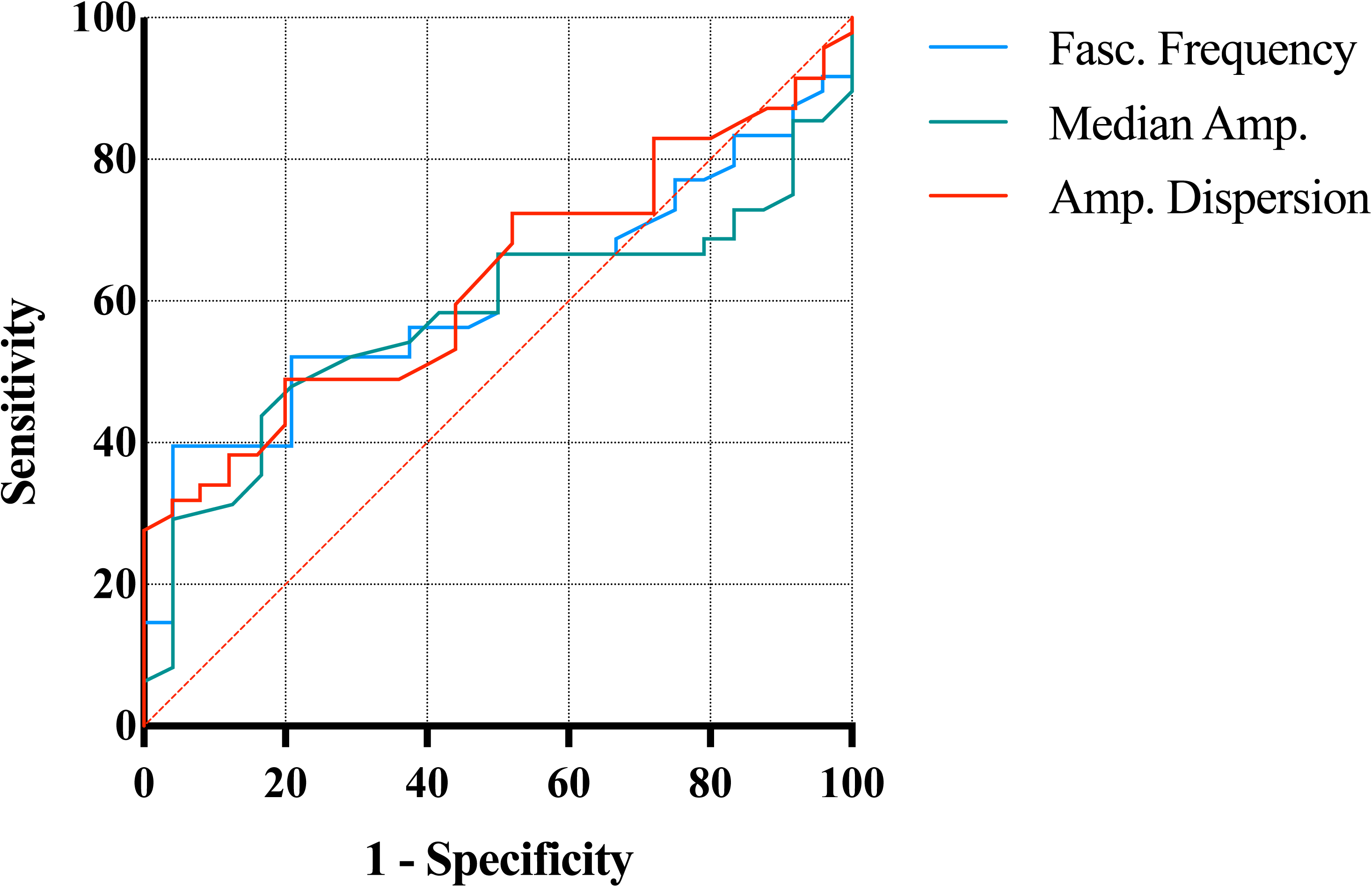
Assessment of the diagnostic utility of fasciculation potential parameters recorded in the medial gastrocnemius in ALS. ROC analysis, after dichotomising outcomes by combining the neurological and healthy control groups (1= ALS, 0 = not ALS), demonstrated areas under the curve of 0.61, 0.58, and 0.63 for fasciculation frequency, fasciculation amplitude, and fasciculation amplitude dispersion, respectively, each value indicative of a poor diagnostic instrument.

### 3.3 Prognostic

#### 3.3.1 Association of fasciculation potential parameters with the prognosis of ALS

Median values (with IQR) across the cohort for FP frequency, FP median amplitude, and FP amplitude dispersion were 38.5 FP/min (10.3-77.6), 24.9 µV (21.3-45.2), and 24.3 µV (14.8-37.9), respectively. Log-rank analysis of the Kaplan-Meier curves for our *predicted* survival showed no statistically significant differences for any of the muscle FP parameters, as seen in *Fig. 3*. However, there was a clear divergence between the dichotomized values of FP median amplitude, therefore this parameter was chosen for Cox proportional hazards analysis. FP median amplitude was associated with a hazard ratio of 1.003 (0.97 – 1.03, p = 0.83) after accounting for bilateral recordings in our dataset.

**Fig. 3:**
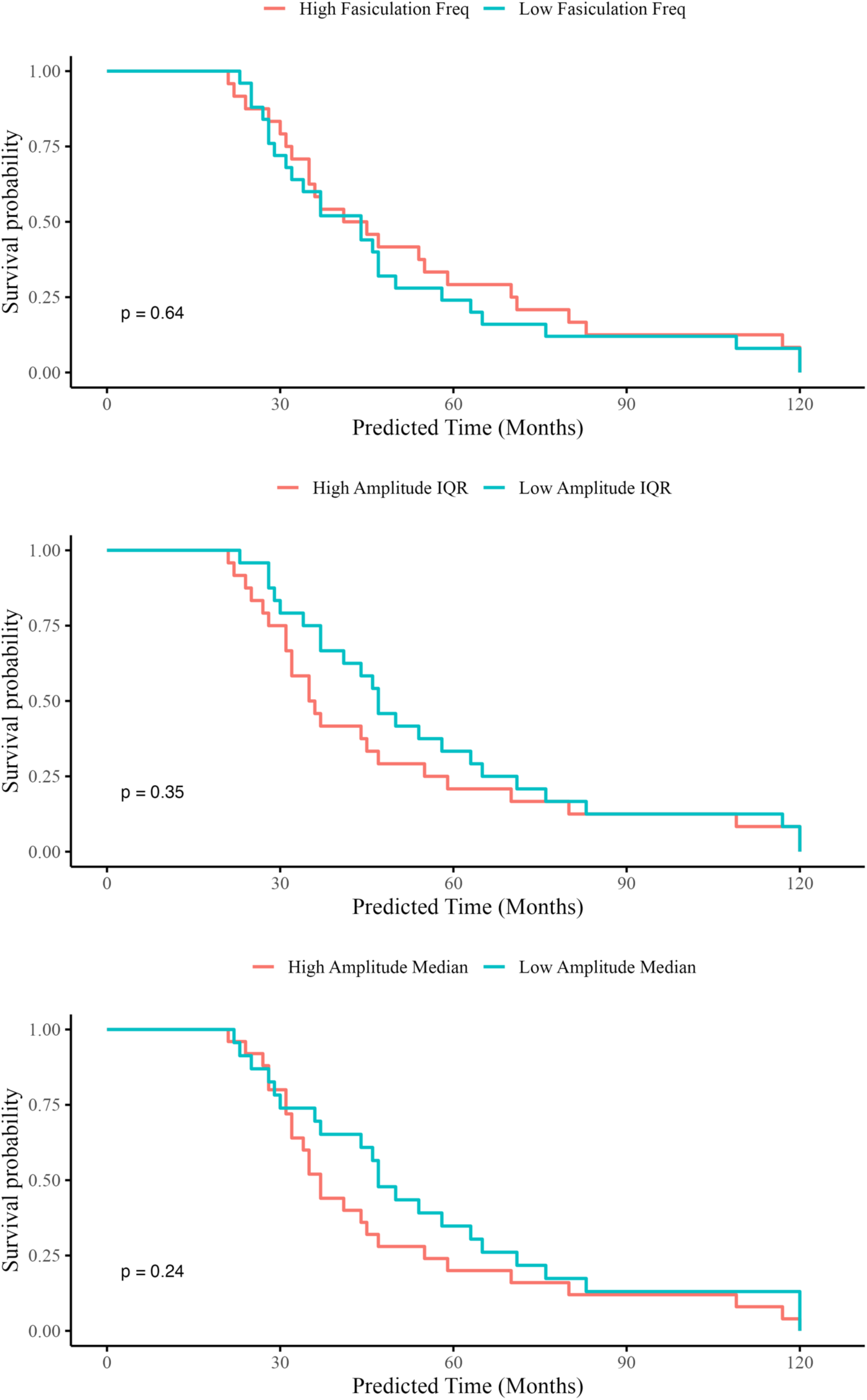
A Kaplan-Meier analysis using real-world, observed survival times from a retrospective cohort of patients with ALS. It involved the analysis of three FP parameters: frequency, median amplitude, and amplitude dispersion. Dichotomization into high and low groups occurred at the median value.

Similarly, as seen in *Fig. 4*, there were no statistically significant differences in the Kaplan-Meier curves of fasciculations in the *observed* survival dataset (observed survival data was recorded for 21 patients, with 28 surviving patients censored from the analysis). Similarly, while not significant, there were clear divergences in dichotomized values for FP frequency and FP median amplitude, thus they were implemented in a Cox model. FP frequency and FP median amplitude were associated with hazard ratios of 1.00 (0.99-1.01, p = 0.96) and 1.00 (0.98-1.03, p = 0.97), respectively.

**Fig. 4:**
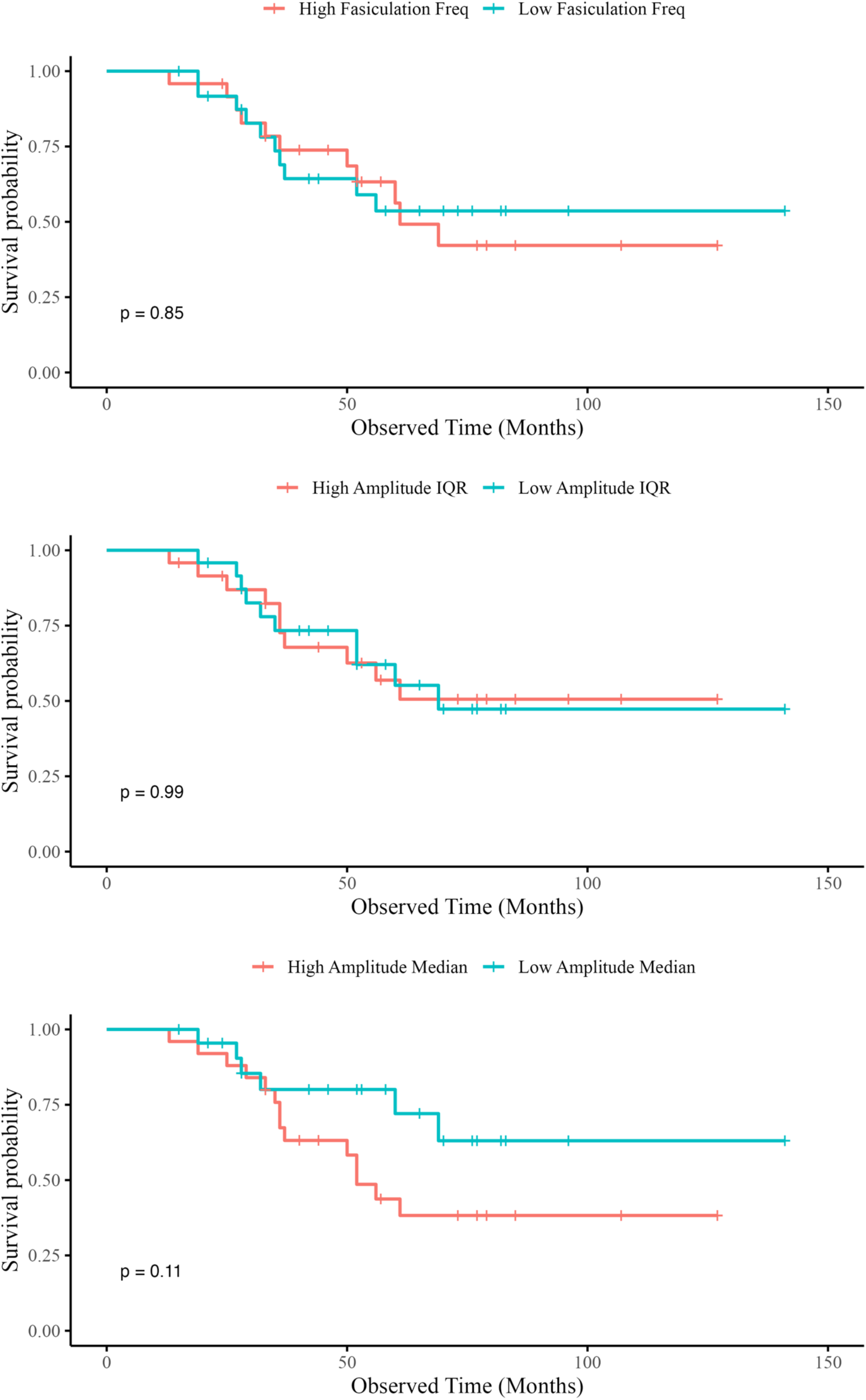
A Kaplan-Meier analysis using a prospective, predicted patient survival using the ENCALS Survival Model. It involved the analysis of three FP parameters: frequency, median amplitude, and amplitude dispersion. Dichotomization into high and low groups occurred at the median value.

## 4. Discussion

Fasciculations are a hallmark clinical feature of ALS and are reflective of early neuronal dysfunction. Therefore, their measurement and interpretation have the potential to elucidate the processes of neurodegeneration in the early stages of the disease course (Bashford et al., 2019) and expedite diagnosis and prognosis. Despite the evident potential of HDSEMG as a diagnostic or prognostic tool in ALS, it is currently not validated for use in the clinic and remains confined to the research setting. ALS diagnosis and prognosis require an integrative approach, making use of clinical history, physical examination and electrodiagnostic tests (Goutman et al., 2022). Since fasciculation assessment in MG performed poorly as a diagnostic and prognostic tool in isolation, we could not justify planned combinatorial analysis with the BB data. However, this study reinforces a statistical framework for the future inclusion of multiple muscles as part of a clinically relevant body-map assessment. Future work is planned from our group, which will focus on the assessment of muscles known to become weak early in the disease course, such as first dorsal interosseous, abductor pollicis brevis, and tibialis anterior.

### 4.1 Poor diagnostic utility of gastrocnemius HDSEMG data

Our results show that HDSEMG recording in the MG fails to add any additional diagnostic insight above that already provided by recordings in the BB (Tamborska et al., 2020). This finding is corroborated by previous comparisons of gastrocnemius FP parameters between ALS and neurological controls, showing similar spatial distribution, intensity (the number of FP pairs), amplitude, duration, and number of phases (FP repeats within 10-110 ms) between groups (Van Der Heijden et al., 1994, Kleine et al., 2012, Sleutjes et al., 2016). Fermont showed that fasciculations occur frequently in the lower leg muscles of healthy people and may be temporarily increased by physical exercise (Fermont et al., 2010), with the theory being that exercise may induce this increase *via* metabolic stress. The gastrocnemius has also shown more diurnal variation in FP frequency than the BB (Bashford et al., 2020a), suggesting values recorded in the gastrocnemius may be more sensitive to the exact timing of HDSEMG recording than in BB. Furthermore, fasciculations in ALS patients have been shown to occur more often in proximal than in distal muscles, and markedly more frequently in the upper than in the lower limbs (Krarup, 2011), further promoting the use of BB or other upper limb muscles in the future assessment of the utility of HDSEMG in ALS.

The limited diagnostic utility of the MG HDSEMG recordings may also be explained by the differences in disease stage between the participants recruited into this study. It has previously been proposed that FP frequency may increase in the early stages of ALS until muscle weakness occurs, after which it subsequently declines (Tamborska et al., 2020). This initial increase may be attributed to the progression of axonal membrane abnormality in the affected neurons (Mills, 2010), whereas the subsequent fall may be attributed to shrinkage of the MU pool and saturation (or reversal) of the relative hyperexcitability of the surviving MUs (Bashford et al., 2020c). Considering this, the time point at which each patient undergoes HDSEMG recording may bear an influence on their FP parameter values, as they may be at different stages of their disease course and exhibiting varying stages of neuropathophysiology. Furthermore, the fascicle architecture of the two muscles may have an impact on their relative sampling of MUs. In comparison to the parallel architecture of the BB, the fascicles of the gastrocnemius have a pennate architecture with spatially localised MU territories (Vieira et al., 2011). So, the extent to which the position of the electrode grid influences which MUs are within the detection volume may be greater in the gastrocnemius than in the BB (where the majority of MUs would run beneath the grid). Furthermore, variability in the data analyses between studies e.g., differences in methods of determining FP discharge frequency (De Carvalho and Swash, 2016, Noto et al., 2018), may have a significant influence on studies’ results and their subsequent interpretation.

### 4.2 Poor prognostic utility of gastrocnemius HDSEMG data

Similarly, our study has shown that muscle FP parameters obtained from the MG muscle are not useful at prognosticating patients with ALS. This result was mirrored using both predicted survival values from the ENCALS Survival Model and real-world observed values.

While our findings are in contradiction with a previous study which focused on BB (Wannop et al., 2021), there are several plausible explanations. This is the first study aimed solely at the prognostic utility of the MG muscle. When examined histologically, the gastrocnemius muscle is comprised mainly of fast-twitch muscle fibres in comparison to the roughly equal ratio of fast twitch to slow twitch found in the BB muscle (Silva Cornachione et al., 2011, Srinivasan et al., 2007). This nuance is important as ALS is known to preferentially affect fast-twitch muscle fibres (Atkin et al., 2005). Therefore, as muscle FP parameters are known to fall with disease progression (Bashford et al., 2020c), one may speculate that the disease may progress rapidly in the gastrocnemius and may only prove to be a useful measurement much earlier in the disease course. When examining our patient population, HDSEMG recordings were not performed at the time of symptom onset, with the interquartile range 19-34 months after symptom onset. Therefore, it is possible to speculate that the clinical utility of the MG muscle may have been lost long before recordings were taken, and future studies should aim to obtain recordings as soon as a clinical suspicion of ALS arises.

Additionally, we chose a different approach for expressing our fasciculation measures than those employed by Wannop *et al*. (Wannop et al., 2021). Where our study used single-timepoint, ‘raw’ recordings, Wannop *et al*. (Wannop et al., 2021) divided these values by months since symptom onset. We decided against implementing this approach in our study. Firstly, as the ENCALS Survival Model uses time variables to produce a predicted survival time, we wanted to avoid spurious correlations, whereby longer prognoses were incorrectly associated with FP parameters (Bashford et al., 2020c). Secondly, there is a weaker relationship between disease progression and change in fasciculation parameters in the MG muscle in comparison to the BB muscle. In particular, weak gastrocnemius muscles show little to no loss of FP frequency as the disease progresses (Bashford et al., 2020c).

Through investigating the prognostic utility of the MG muscle, we have shown that HDSEMG recordings have little prognostic utility in the middle-to-late disease course. We have speculated that HDSEMG recordings close to the time of symptom onset may prove to be the most reliable and we hope that the findings from this study can support prospective data collection in other, more reliable muscles.

### 4.3 Future directions of study

This study has shown that the analysis of fasciculations recorded in the MG following symptom onset does not show diagnostic or prognostic utility in ALS. However, HDSEMG may provide a wide range of benefits for ALS patients and their families, as the non-invasive nature of the recording allows for longer recording periods and greater patient comfort (Mateen et al., 2008, Howard and Murray, 1992, Bashford et al., 2019). It is also capable of achieving greater spatial resolution and muscle area coverage than is possible with needle EMG, allowing for more precise anatomical mapping of fasciculations (Drost et al., 2007). Due to this greater coverage, there is also less reliance on the reproduction of needle position in serial recordings, making HDSEMG more suitable for the collection of longitudinal data (Benatar et al., 2016, Bashford et al., 2020b). This is essential for capturing the dynamic changes that occur in ALS due to the inexorable decline of motor neurons (Bashford et al., 2020b). Perhaps the most important benefit of this technique is its ease of application and potential for use outside of the neurophysiology lab. Its use by non-clinicians in the community could provide greater relief to patients and their families as well as benefiting medical research by analysing the disease at a pre-symptomatic stage (Bashford et al., 2020b).

Therefore, further research into the application of HDSEMG in the diagnosis and prognosis of neuromuscular disease is required and should focus on the BB or other upper limb muscles at an earlier stage in the disease course. Due to the presence of fasciculations in both ALS patients and controls, HDSEMG data should be considered in the context of other clinical factors, such as the distribution of fasciculations, other neurological symptoms, and a clinical suspicion of ALS (Noto et al., 2018, Mills, 2010, Drost et al., 2007, de Carvalho et al., 2017, De Carvalho et al., 2008). A combinatorial approach may be most appropriate for this, using tools such as electrical impedance myography, ultrasound, and magnetic resonance imaging to monitor clinical, electrophysiological, biochemical, and imaging biomarkers simultaneously.

Large numbers of patients across the spectrum of ALS phenotypes should also be monitored in longitudinal studies with post-mortem validation, including prospective multi-centre cohort analyses, to develop reliable and sensitive biomarkers. The greatest diagnostic challenge appears to be in differentiating between ALS and neurological controls. So, a greater range of ALS mimics e.g., spinal muscular atrophy, MMN and spinobulbar muscular atrophy (Kennedy’s disease) should be included in neurological control groups.

## 5. Conclusion

The present study has been successful in producing an analytical framework that may be reproduced for the diagnostic and prognostic evaluation of HDSEMG in novel muscle groups. While our results in MG were negative, we believe this study supports the argument for early HDSEMG recordings across a broader range of clinically relevant muscles in patients with ALS, as it is in these early stages of the disease that fasciculations may prove to be the most valuable.

## Data Availability

All data produced in the present study are available upon reasonable request to the authors.

## 6. Funding

JB acknowledges funding from the Medical Research Council and Motor Neurone Disease Association (Lady Edith Wolfson Clinical Research Training Fellowship; MR/P000983/1), Sattaripour Charitable Foundation, UK Dementia Research Institute, and the National Institute for Health and Care Research (Academic Clinical Lectureship program). DP, UM, and CC contributed during their MSc qualifications in Clinical Neuroscience. R.I.’s input represents independent research supported by the NIHR BioResource Centre Maudsley at South London and Maudsley NHS Foundation Trust (SLaM) & Institute of Psychiatry, Psychology and Neuroscience (IoPPN), King’s College London. RM acknowledges funding from the Faculty of Life Sciences and Medicine, King’s College London.

## 7. Competing interests

The authors declare no relevant competing interests.

## List of abbreviations

ALS: amyotrophic lateral sclerosis
UMN: upper motor neuron
LMN: lower motor neuron
HDSEMG: high-density surface electromyography
MU: motor unit
FP: fasciculation potential
EMG: electromyography
BB: biceps brachii
ROC: receiver operating characteristic
CHAID: chi squared automatic interaction detection
CI: confidence interval
MG: medial gastrocnemius
ENCALS: European Network to Cure ALS
BFS: benign fasciculation syndrome
PN: peripheral neuropathy
MMN: multifocal motor neuropathy
ALSFRS-R: revised amyotrophic lateral sclerosis functional rating scale
SPiQE: surface potential quantification engine
IQR: interquartile range
ANOVA: analysis of variance.

## Notes

### Competing Interest Statement

The authors have declared no competing interest.

### Funding Statement

JB acknowledges funding from
Medical Research Council and Motor Neurone Disease Association (Lady Edith Wolfson Clinical Research Training Fellowship; MR/P000983/1)
Sattaripour Charitable Foundation
UK Dementia Research Institute
National Institute for Health and Care Research (Academic Clinical Lectureship program).

